# SIR analytical model applied to 2020 Covid-19 data in Italy

**DOI:** 10.1101/2022.11.09.22282122

**Authors:** Roberto Simeone

## Abstract

A simple analytical Susceptible-Infectious-Recovered (SIR) model without vital dynamics is developed from basic assumptions. Based on the data available for the spread of the Covid 19 virus in Italy in 2020, the characteristic parameters of the model are estimated.

## 1 SIR model

When a process can be represented by the temporal evolution of some variables that change their belonging from one category to another, if the interactions are not too complex, it might be useful to develop a ‘compartmentalized’ or ‘compartmnental model’.

In the case of the spread of infectious diseases, a simple model called SIR (Susceptible, Infectious, Removed) is often used^1^. The three letters correspond to the three compartments into which the population can be schematically divided in the event of a pandemic.

If at any given moment a set of people is found with infected individuals who are spreading a disease, that set can be divided into three categories (or compartments):

- 1. Susceptible : healthy individuals who can be infected by proximity or contact with infected individuals;
- 2. Infected : individuals who have the viral vector and who can infect Susceptibles;
- 3. Removed : individuals who have had the disease and are no longer contagious either by death or recovery. In our case we will simplify considering the recovered also immune in the sense that they cannot be infected again.

From the day the disease appears in the population (zero day), the disease will spread from infected people to susceptible people due to interactions between them or their sufficient physical proximity. On the other hand, after a certain period of time, infected people will become unable to spread the virus (thus passing into category R) either by having acquired immunity or by having adopted prescriptions and social restrictions that drastically reduce interactions (e.g. protective devices and lockdown).

If the diffusion is much faster than the vital dynamics of the affected population, in the sense that during the observed period the variations due to births and deaths can be neglected, the total population, in the model, can be considered constant. In this case we will consider the vital dynamic negligible

In the simple case of homogeneous distribution of the three SIR categories in the population, it can be assumed that the number of Susceptibles that becomes Infected in the unit of time (*α*_*SI*_) depends on the average number of Susceptibles that are contacted by an Infected each day(*r*) multiplied by the probability (*p*) of transmission for each contact.

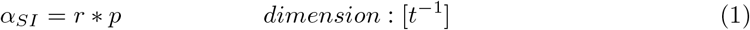

On the other hand, the average transfer rate from Infected to Removed (*α*_*IR*_) is basically the reciprocal of the average period in which the Infected are contagious, before moving on to the Recovered category. Its value depends on many factors, beyond the average duration of the illness, such as the adoption of protective equipment and social limitations.Each infected individual in the period in which it is contagious will produce on average a number of new infected equal to the average number of contacts per Suceptible per day multiplied the duration of the period in which the infected is contagious, that is to say:

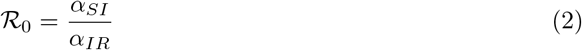

ℛ_0_ it is called Basic Reproduction Number^2^ and represents the average number of new infected produced by each infected in its infectious period.

Let us now consider the variation over time in the number of individuals belonging to the three categories, considering for simplicity that the total number *N* does not change in the period concerned.

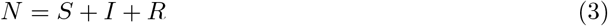

where :

- *S*(*t*) is the number of individuals belonging to the Susceptible category;
- *I*(*t*) is the number of individuals belonging to the Infected category;
- *R*(*t*) is the number of individuals belonging to the Removed category;
- *N* = *S*(*t*) + *I*(*t*) + *R*(*t*) is the total number of individuals that does not change over time.

From the moment *t* = 0 in which infected individuals are present, the number of Susceptibles decreases contributing to the corresponding increase in the number of Infected.

On the other hand, the number of Infected, while it is increased by the decrease of Susceptible, is reduced by the number of infected who have lost the ability to infect, thus moving into the Removed category.

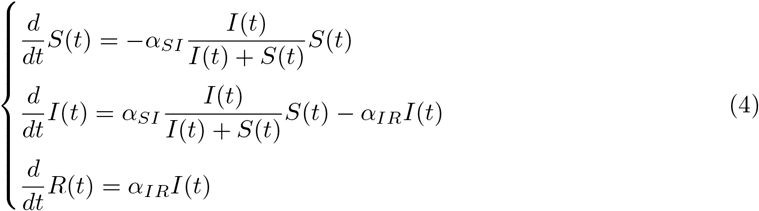

In this case we assume that the transmission rate *α*_*SI*_ depends on the average number of contacts per person related to *I* and *S*, then we assume that the variation of *S*(*t*) over time is the product between the transmission rate (between *S* and *I*), the fraction of Infected on the population not yet Recovered, and the number of Susceptibles.

It should be noted that in SIR model is commonly used the average number of contacts per person per day, then the *I/N* ratio between infected and the total population is used instead the *I/*(*I* + *S*) used here.

To simplify the reading we will use, according to Newton’s notation, the point above the variable to indicate the time derivative and we will indicate the SIR variables with the indices 1,2,3 without always explicitly indicating the time dependence.

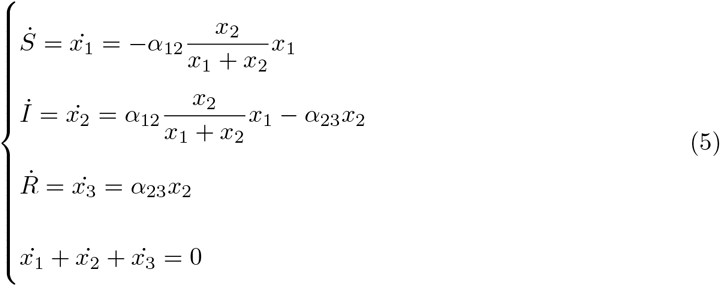

## 2 analytical solution

The first two equations of 5 can be written as:

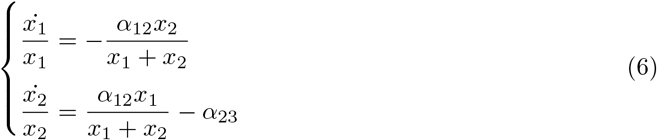

Subtracting the two equations of 6 we obtain:

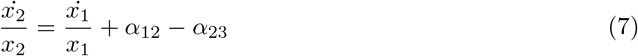

Considering that 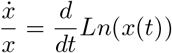, integrating the 7 we obtain:

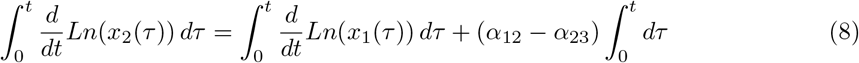

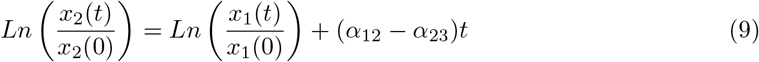

Raising both members to power we have:

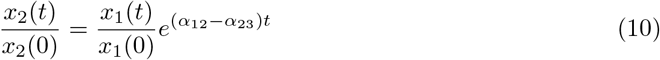

The 10 thus becomes:

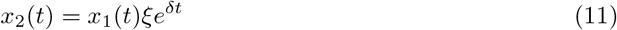

where:

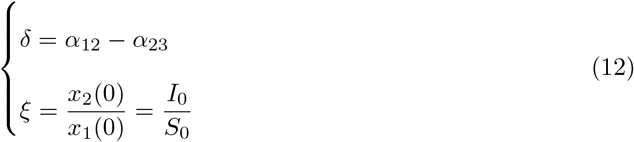

*S*_0_ = *x*_1_(0) ; *I*_0_ = *x*_2_(0) represent the initial values of the number of Susceptible and Infected individuals.

At the initial instant, of course, there are no Recovered, therefore: *R*_0_ = *x*_3_(0) = 0.

Substituting 11 in 6 we obtain:

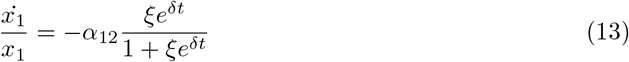

Two derivatives can be recognized in 13:

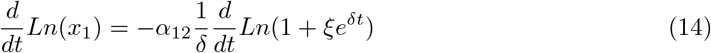

Integrating the 14 we obtain:

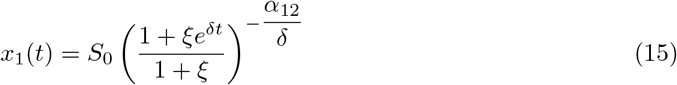

Substituting 15 into 11 we obtain:

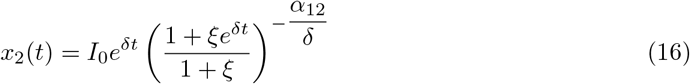

At this point, since we are neglecting the vital dynamic, *N* = *x*_1_ + *x*_2_ + *x*_3_. so:

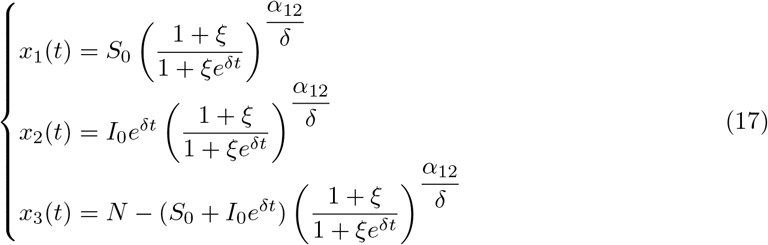

### 2.1 Fitting the curve *I*(*t*)

Remembering that *I*(*t*) = *x*_2_(*t*) for the 17 we can write:

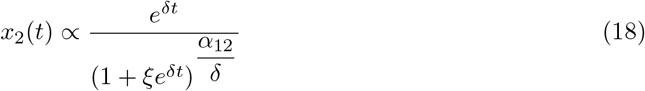

It can be assumed that the curve initially grows as *e*^*δt*^, while for *t* → +∞ in 18 the unit constant in the denominator can be neglected and we have that:

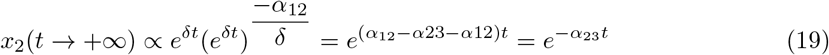

Summarizing, having available the data relating to *I*(*t*), if these represent in a sufficiently clear way the initial and final exponential trend (there are not too many wrong data or outliers), then the values of *δ*and *α*_23_ can be obtained by fitting the initial growth 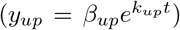 and the decreasing tail 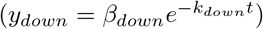, also getting *α* ^12^ = *δ* + *α* ^23^.

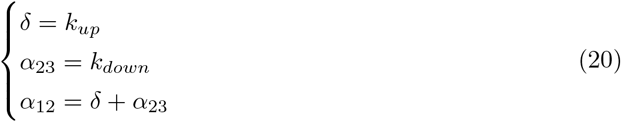

The fitting can be easily done using the linear Least Square method applied to the logarithm of the data.

### 2.2 The maximum of *I*(*t*)

Referring to the second equation in 6 we can write:

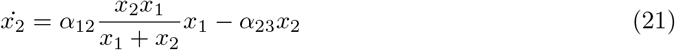

Substituting the explicit functions of *x*_1_ and *x*_2_ according to 17 and collecting the terms we can write:

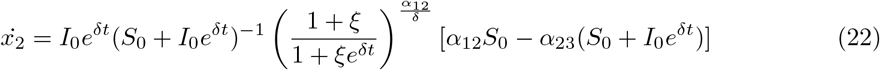

The time derivative of *x*_2_ vanishes when the content in the square bracket is zero:

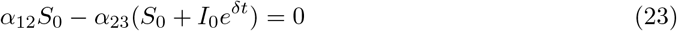

or also:

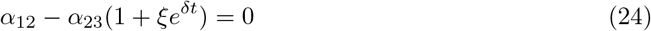

The value of *t* that cancels this term is:

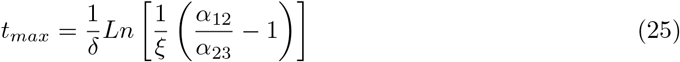

It can be noted that the existence of a maximum implies that 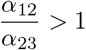, so it must be *α* ^12^ > *α* ^23^.

The 24 can also be written:

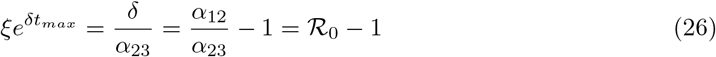

Knowing *δ, α*_23_ and *t*_*max*_ we can therefore also obtain:

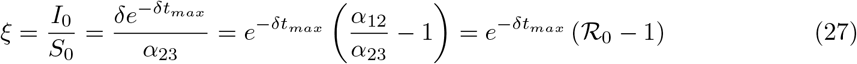

Considering that for the 17 we can write *x*_2_(*t*) = *ξe*^*δt*^*x*_1_(*t*), in the case of *t* = *t*_*max*_ for 26 we also obtain the ratio, at time *t*_*max*_, between the number of Susceptible and that of Infected:

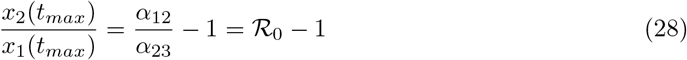

## 3 Available data

We will apply the model to the data referring to the new cases of Covid 19 detected in Italy in the period 2020/2/24-2020/7/22, registered by the Italian Department of Civil Protection and available on the site indicated in the footnote. ^3^.

It should be noted that in the period from 2020-03-11 to 2020-05-04 (lockdown period) strong social limitations were adopted in addition to the obligations to adopt protective devices.

**Table 1.**
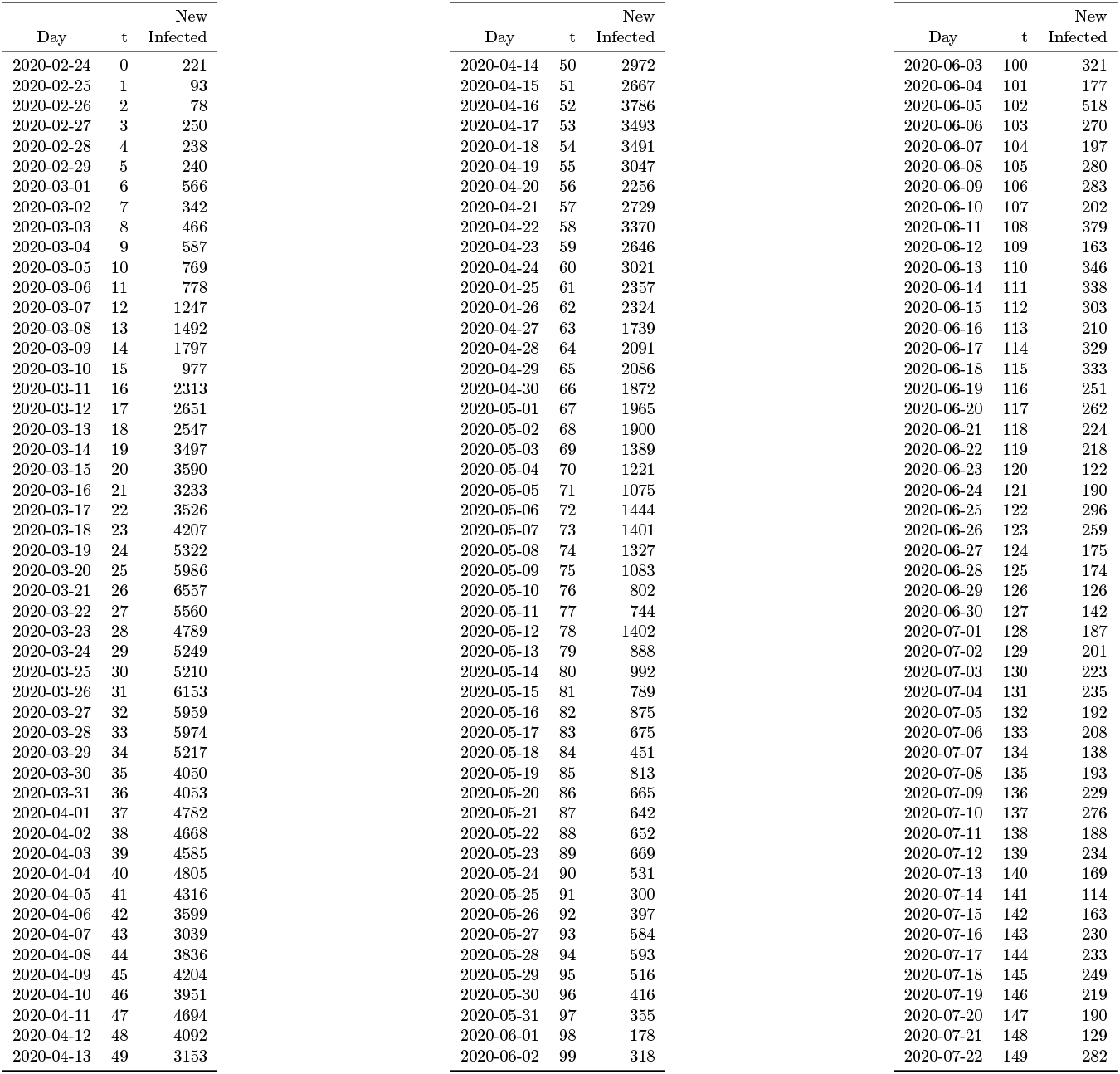
New Covid 19 positives in the period 2020/2/24 - 2020/7/22 recorded by the Italian Department of Civil Protection.

## 4 Exponential fitting and model

The data in fig 1 can be represented for the initial phase by an exponential growth 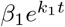 and, in the final phase, by an exponential decrease 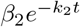 con *k*_1_, *k*_2_ > 0 (see fig. 2).

**Figure 1:**
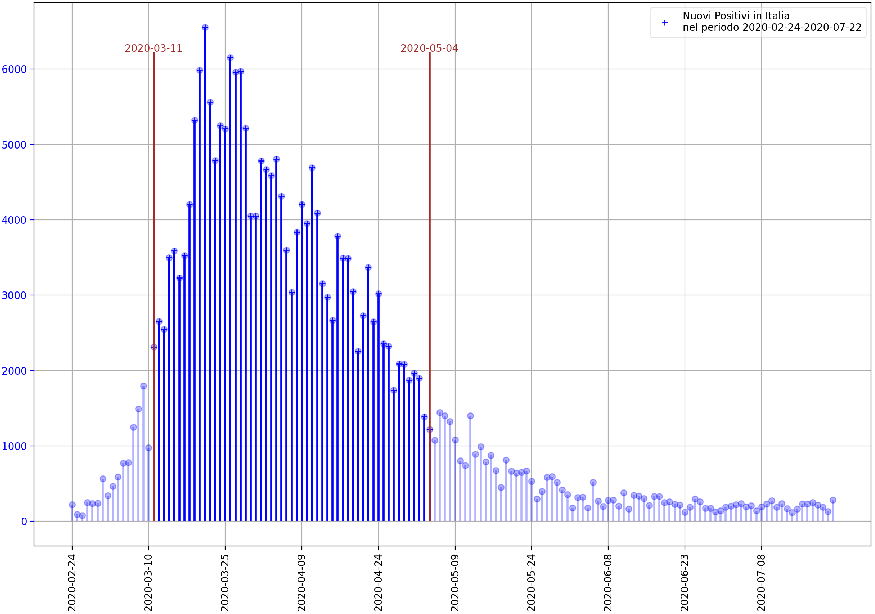
Trend of new Covid 19 positives in Italy in the period 2020/2/24 - 2020/7/22 recorded by the Italian Department of Civil Protection. The ‘lockdown’ period is higligted in the figure.

**Figure 2:**
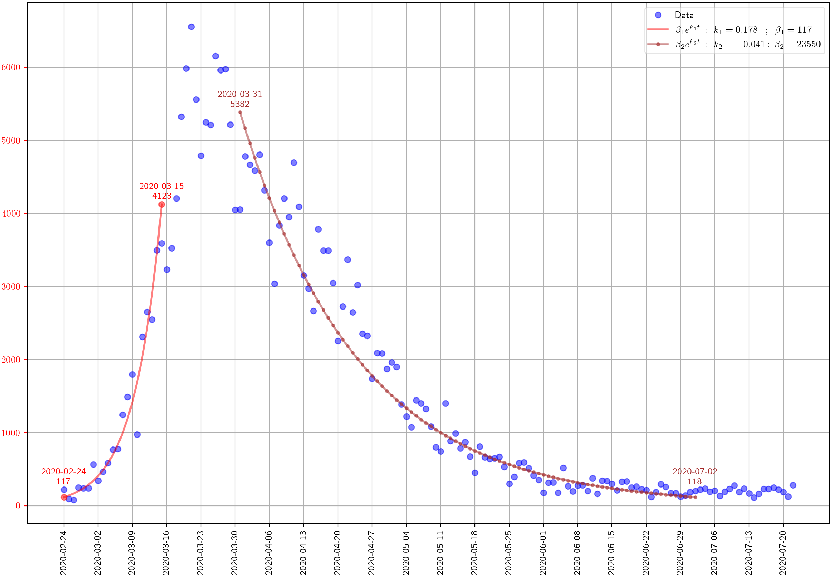
Fitting the data of the new positives in fig. 1 with the exponential βe^kt^

As stated in par. 2.1 and 2.2 we have:

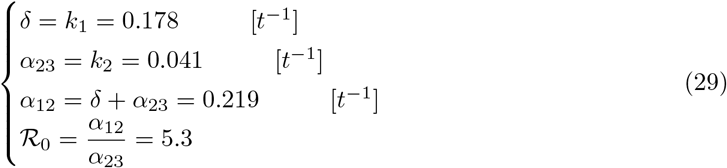

Since from the available data (see tab. 1) it can be deduced that the maximum of the *I*_*t*_ curve is between 25^*th*^ and 33^*th*^ day, setting *t*_*max*_ = 30, for 27 we obtain *ξ*:

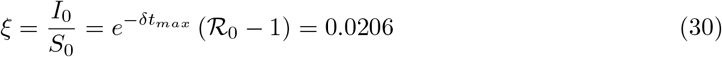

This implies that if *I*_0_ = 221 new cases are registered at the initial moment, it is expected that there are *S*_0_ = *I*_0_*/ξ* = 10728 individuals Susceptibles involved in the modeled process (see fig. 3).

**Figure 3:**
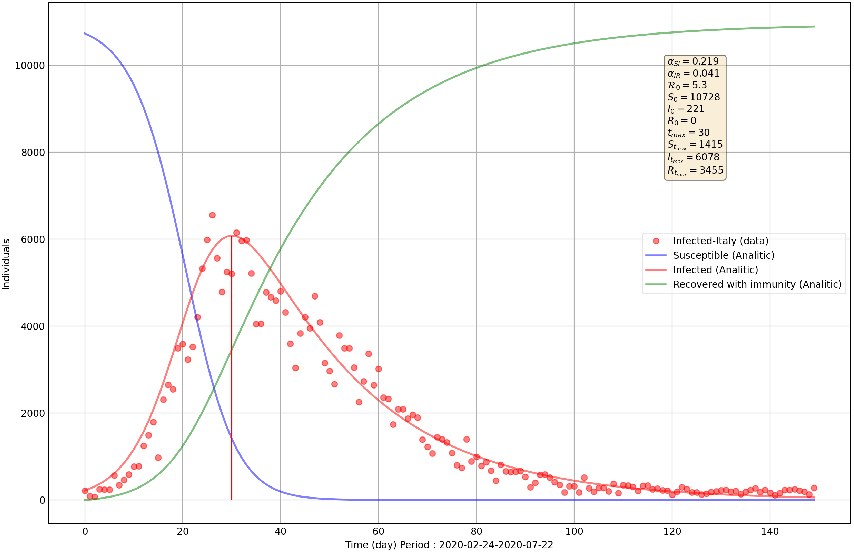
SIR model based on the fitting in fig.2.

With these assumptions it can be estimated that the first case occurred at the time 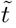, with:

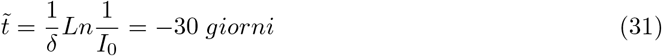

## 5 Conclusions

In this simple model, starting from the *I*(*t*) curve of the new infected, it is possible to easily obtain exponential parameters of growth and decrease to which the infectivity rates (*α*_*SI*_) and recovery rates (*α*_*IR*_). are correlated.

By identifying also the time corresponding to the maximum of the curve, it is possible to estimate the total number of individuals involved in the diffusion process.

The application of the model to the data of the new infected in Italy in the period 2020/2/24-2020/7/22 led to the following results:

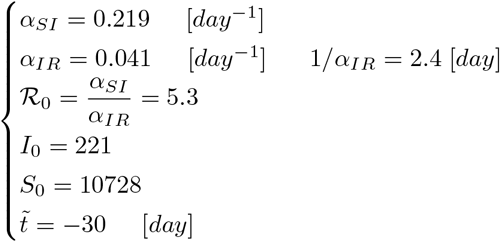

Where :

- *α*_*SI*_ : is the average number of contacts per day between Infected and Susceptibles;
- *α*_*IR*_ : is the reciprocal of the mean duration of infection;
- ℛ_0_ : is the basic reproduction number, or the average number of new Infected produced by one infected individual during his contagious period;
- *I*_0_ : is the number of new infected cases registered at *t* = 0;
- *S*_0_ : is the estimated number of susceptible individuals; involved in the spread process at *t* = 0;
- 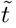 : is the estimated number of days before *t* = 0 when the first infected individual appeared.

The model offers indicative results in the hypothesis of a group of people involved in the spread of the virus in which the three categories *S, I, R* are homogeneously mixed and the estimated parameters must be considered as averages over the observed period.

## Data Availability

All data produced are available online at the Italian Department of Civil Protection : https://mappe.protezionecivile.gov.it/it/mappe-emergenze/mappe-coronavirus/situazione-desktop

https://github.com/pcm-dpc/COVID-19/blob/master/dati-regioni/dpc-covid19-ita-regioni.csv

## A Appendix

### A.1 Application to data relating to the Lombardy region

**Figure 4:**
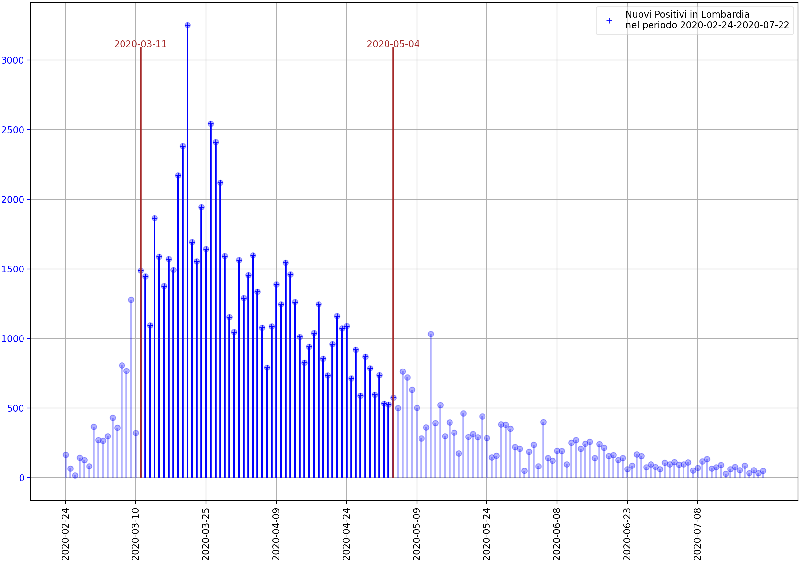
Trend of new Covid 19 positives related to the Lombardy region in the period 2020/2/24 - 2020/7/22 recorded by the Italian Department of Civil Protection.The ‘lockdown’ period is higligted in the figure.

**Figure 5:**
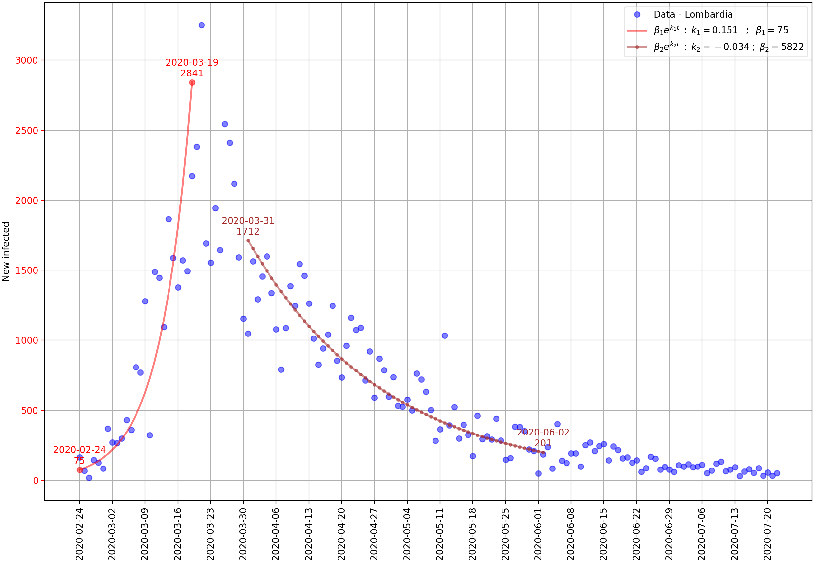
Fitting the data of the new positives in fig. 1 with the exponential βe^kt^

**Figure 6:**
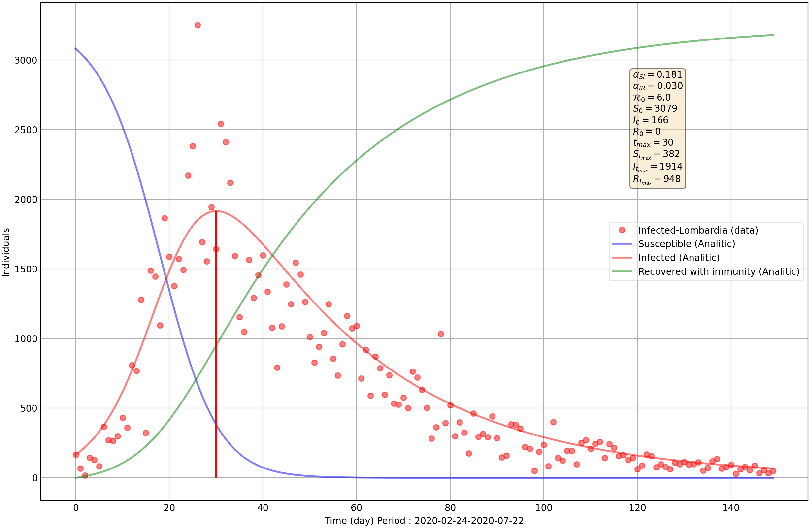
SIR model based on the fitting in fig.5.

### A.2 Application to data relating to the Lazio region

**Figure 7:**
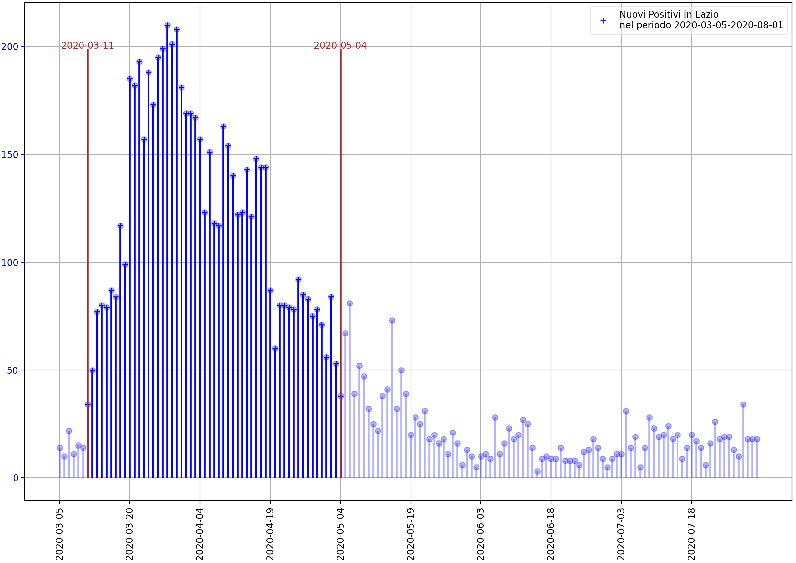
Trend of new Covid 19 positives related to the Lombardy region in the period 2020/2/24 - 2020/7/22 recorded by the Italian Department of Civil Protection.The ‘lockdown’ period is higligted in the figure.

**Figure 8:**
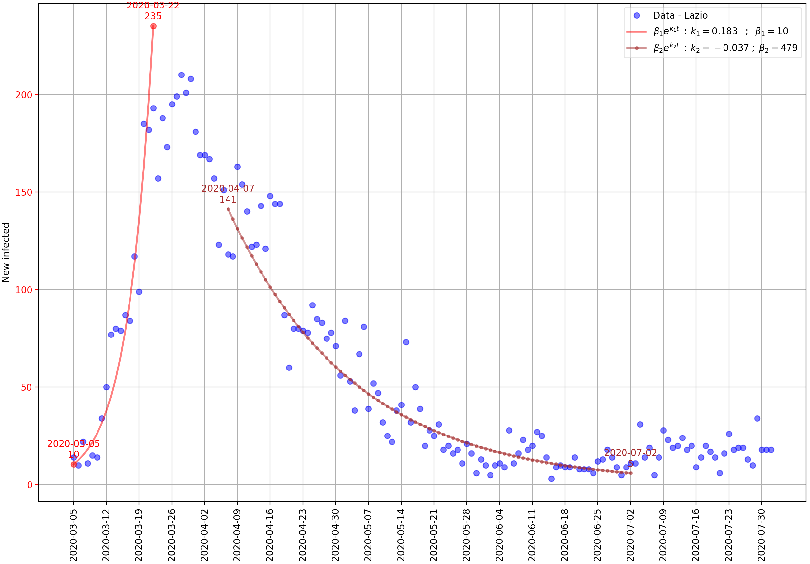
Fitting the data of the new positives in fig. 7 with the exponential βe^kt^

**Figure 9:**
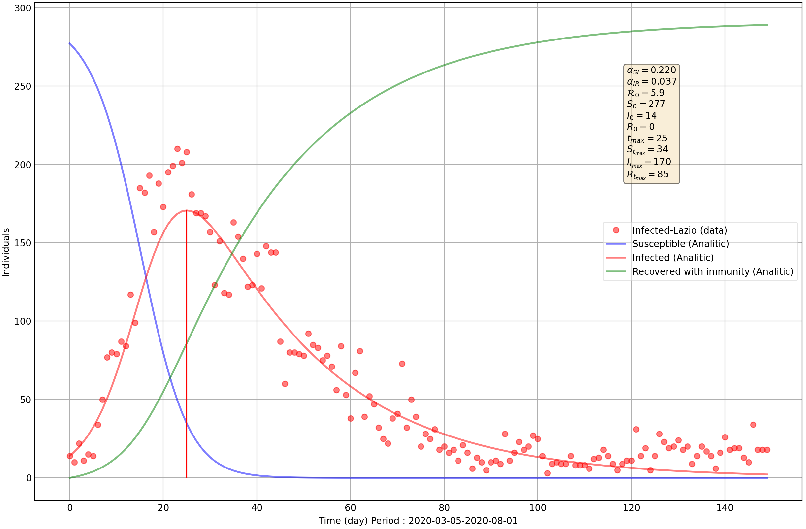
SIR model based on the fitting in fig.8.

### A.3 Summary of the estimated results

| Region | $\alpha_{SI}$<br>$day^{-1}$ | $\alpha_{IR}$<br>$day^{-1}$ | $\mathcal{R}_0$ | People Involved<br>$S(0) + I(0)$ | Start of<br>observed period | First case<br>estimated at | Maximum I(t)<br>estimated at |
| --- | --- | --- | --- | --- | --- | --- | --- |
| Italy | 0.219 | 0.041 | 5.3 | 1728+221 | 2020-02-24 | 2020-01-24 | 2020-03-25 |
| Lombardia | 0.181 | 0.030 | 6.0 | 3079+166 | 2020-02-24 | 2020-01-21 | 2020-03-25 |
| Lazio | 0.220 | 0.037 | 5.9 | 277+14 | 2020-03-05 | 2020-02-19 | 2020-03-30 |

## Footnotes

1 https://en.wikipedia.org/wiki/Compartmental_models_in_epidemiology

2 https://en.wikipedia.org/wiki/Basic_reproduction_number

3 Dipartimento della Protezione Civile COVID-19 Italia - Monitoraggio della situazione https://mappe.protezionecivile.gov.it/it/mappe-emergenze/mappe-coronavirus/situazione-desktop https://github.com/pcm-dpc/COVID-19/blob/master/dati-regioni/dpc-covid19-ita-regioni.csv

## Notes

### Competing Interest Statement

The authors have declared no competing interest.

### Funding Statement

This study did not receive any funding

